# Functional imaging to guide network-based TMS treatments: toward a tailored medicine approach in Alzheimer’s disease

**DOI:** 10.1101/2020.12.22.20248694

**Authors:** Chiara Bagattini, Debora Brignani, Sonia Bonnì, Roberto Gasparotti, Michela Pievani

## Abstract

**INTRODUCTION:** Transcranial magnetic stimulation (TMS) has gained increasing attention as a potential therapeutic strategy in Alzheimer’s disease (AD). Among factors determining a clinical response, the choice of the stimulation site represents a key point. In this proof of concept study, we prove the feasibility of a tailored TMS targeting approach for AD, which stems from a network-based perspective. Based on functional imaging, the procedure allows to extract individual optimal targets meanwhile accounting for functional variability.

**METHODS:** Single-subject resting-state fMRI was used to extract individual target coordinates of two networks primarily affected in AD, the default mode and the fronto-parietal network. The localization of these targets was compared to that of traditional group-level approaches and tested against varying degrees of TMS focality.

**RESULTS:** The distance between individual fMRI-derived coordinates and traditionally-defined targets was significant for a focality <12mm, but not for >20mm. Comparison with anatomical labels confirmed a lack of 1:1 correspondence between anatomical and functional targets.

**DISCUSSION:** The proposed network-based fMRI-guided TMS approach allows targeting disorder-specific networks meanwhile accounting for inter-individual functional variability in Alzheimer’s disease. This approach might represent a step toward tailored TMS interventions for AD.

## 1. Introduction

Through the repeated delivery of short-lived magnetic fields over the scalp, repetitive transcranial magnetic stimulation (rTMS) is able to induce long-lasting changes of cortical excitability, which resemble long-term potentiation or long-term depression-like mechanisms, depending on the stimulation parameters (Wassermann et al., 2008). Robust evidence proves that TMS acts beyond the site of stimulation, affecting the connectivity of the stimulated networks (Fox et al., 2012b; Ruff et al., 2009; Siebner et al., 2009), thus rising considerable interest for its therapeutic application across a range of diseases with distributed network pathology (Fox et al., 2012a; Lefaucheur et al., 2014). In this scenario, rTMS has gained increasing attention as a potential treatment also in the battle against Alzheimer’s disease (AD) (Weiler et al., 2020), for which the disappointing outcome of recent clinical pharmacological trials highlights the urgent need for effective alternative interventions.

Evidence regarding clinical efficacy of rTMS treatment in AD, however, is feeble and key issues remain before its clinical application (Lefaucheur et al., 2020). Among factors determining the clinical response, two crucial aspects are represented by the rationale for choosing a given site of stimulation and the procedure to target this region. The majority of previous rTMS studies individuated the target areas through coarse procedures, such as rule of thumb, EEG electrode system, group-averaged coordinates or anatomical landmarks (please refer to Table 1 for an overview of methods adopted in previous studies).

**Table 1.** Summary of the target areas and localization methods adopted in previous rTMS interventions in AD patients.

| Study | Target area(s) | Localization method |
| --- | --- | --- |
| *5-cm rule* |  |  |
| Ahmed et al., 2012 | Left and right DLPFC | 5 cm rostral to optimal site for motor threshold production in the first dorsal interosseous |
| Haffen et al., 2012 | Left DLPFC | 5 cm anterior and parasagittal from the hand area |
| Drumond Marra et al., 2015 | Left DLPFC | 5 cm in a parasagittal plane parallel to the point to maximum stimulation of the short abductor of the thumb |

| *Electrode position(s) according to the International 10-20 EEG System* |  |  |
| --- | --- | --- |
| Zhao et al., 2017 | Left and right parietal and posterior-temporal areas | P3, P4, T5, T6 |
| Alcalá-Lozano et al., 2018 | Broca, Wernicke, right and left DLPFC, right and left pSAC | Left DLPFC: electrode not defined; other regions: localization method not defined |
| Turriziani et al., 2019 | Right DLPFC | F4 |
| Bagattini et al., 2020 | Left DLPFC | F3 |
| *Group-average coordinates (mean Tailarach coordinates)* |  |  |
| Cotelli et al., 2010 | Left DLPFC (BA 8/9) | x=-35, y=24, z=48 |
| Cotelli et al., 2012 | Left IPL | x=-44, y=-51, z=43 |
| *Individual anatomical landmarks* |  |  |
| Bentwich et al., 2011 | Broca, Wernicke, right and left DLPFC, right and left pSAC | Identified by neuroradiologist on individual MRI scans |
| Rabey et al., 2013 | Broca, Wernicke, right and left DLPFC, right and left pSAC | Identified by neuroradiologist on individual MRI scans |
| Rabey and Dobronevsky, 2016 | Broca, Wernicke, right and left DLPFC, right and left pSAC | Not better defined |
| Lee et al., 2016 | Broca, Wernicke, right and left DLPFC, right and left pSAC | Identified by neuroradiologist on individual MRI scans |
| Nguyen et al., 2018 | Broca, Wernicke, right and left prefrontal cortex, right and left parietal cortex | Identified by the Neuronix neuronavigation system based on the individual MRI. |
| Koch et al., 2018 | Precuneus | Individual T1-weighted MRI volumes were used as anatomical reference |
| Sabbagh et al., 2020 | Broca, Wernicke, right and left DLPFC, right and left parietal cortex | Brain regions were marked in individual MRI scan by projecting the relevant brain region onto the scalp |

These approaches, however, do not account for the functional organization of the brain and the synaptic dysfunction affecting specific networks in AD. AD is associated with disruption of two large-scale networks central to cognition, the Default Mode Network (DMN) and the Fronto-Parietal Network (FPN) (Agosta et al., 2012; Pievani et al., 2014). The DMN is medially anchored to the posterior cingulate cortex/precuneus and ventromedial prefrontal cortex, and to the bilateral parietal (inferior parietal lobule – IPL, which include the angular and inferior parietal gyri), temporal (lateral temporal cortex and hippocampi), and frontal cortex (dorsolateral prefrontal cortex – DLPFC, roughly corresponding to the superior frontal gyrus). The FPN includes the bilateral DLPFC (middle frontal gyrus) and parietal (superior parietal gyrus) cortex. Due to their crucial role in modulating cognition in AD, these functional networks might represent valid targets for rTMS treatments in this population. The clinical promise of stimulating AD-core networks such as DMN is demonstrated by a recent study showing an improvement in memory by targeting the precuneus (Koch et al., 2018). Moreover, although some of the previous rTMS studies might have stimulated regions belonging to these networks (i.e., DLPFC node of the FPN, IPL node of the DMN; Lefaucheur et al., 2020), this remains speculative lacking a direct assessment with neuroimaging.

Interestingly, some studies have already used network connectivity to guide TMS target selection in healthy young (Momi et al., 2020; Santarnecchi et al., 2018) and elderly participants (Nilakantan et al., 2019; Wang et al., 2014), as well as in psychiatric patients (Fox et al., 2012a; Hoffman et al., 2007), but not in AD. Given the potential value of tailored network-based rTMS intervention for neurocognitive and psychiatric diseases, here we demonstrate the feasibility of a TMS approach that uses resting-state fMRI to identify and target functionally, patho-physiologically and clinically relevant AD networks at the individual level. This strategy is compared to traditional approaches for target localization.

## 2. Methods

We included 8 AD patients (age: 75.3 years [min 69 – max 80]; 6 females, MMSE: 21.6 [min 18 – max 25]) with a clinical diagnosis of AD (McKhann et al., 2011) recruited between June 2019 and February 2020 at the IRCCS Fatebenefratelli (Brescia, Italy) in the context of a randomized controlled clinical trial (GR-2016-02364718; NCT04263194). The study was approved by the local ethics committee and participants signed a written informed consent.

MRI scans were acquired on a 3T Siemens Skyra scanner equipped with a 64-channels head-neck coil at the Neuroradiology Unit, Spedali Civili Hospital (Brescia, Italy). Multiband accelerated rs-fMRI (TR=1000ms, TE=27ms, flip angle=60°, voxel size = 2.1mm isotropic, 70 slices, 600 volumes) and 3D T1-weighted (TR=2300ms, TE=2ms, flip angle=9°, voxel size = 1mm isotropic, 176 slices) scans were collected. Rs-fMRI data pre-processing was carried out using the FMRIB’s Software Library (FSL, http://www.fmrib.ox.ac.uk/fsl/; Smith et al., 2004) and included removal of the first ten time-points, correction of motion with FLIRT and correction of susceptibility-induced distortions with TOPUP (Andersson et al., 2003). The networks of interest (DMN and FPN) were extracted from individual rs-fMRI scans using independent component analysis (ICA) with Melodic (Beckmann and Smith, 2004; https://fsl.fmrib.ox.ac.uk/fsl/fslwiki/MELODIC). Melodic processing included high-pass temporal filtering (0.01Hz), smoothing with a 4mm FWHM filter, affine transformation of EPI images to native T1 images and nonlinear warping of T1 images to standard MNI space. The number of components was automatically estimated by Melodic. The networks of interest in MNI space were identified using a template matching procedure with published templates (Shirer et al., 2012). To ensure the reliability of the components, Melodic was run 10 times and the spatial map most frequently classified as ‘DMN’ or ‘FPN’ was retained. Spatial maps are expressed as z-scores indicating the degree of activation (versus noise) of each voxel within the component. The selected DMN and FPN spatial maps were then back-transformed to subjects’ native T1 space using Melodic transformations. FSL’s *cluster* routine was used to decompose each network into clusters, which are provided with information on their size, coordinates, and maximum intensity. For each subject, DMN and FPN candidate targets were defined as the peak (local maxima) within the largest cluster located, respectively, in the left IPL and left DLPFC (defined by visual inspection). The local maxima were overlaid onto the native T1 scan and the final target was selected according to the following criteria: (i) location specific to the network of interest (i.e., coordinates falling within the spatial maps of both DMN and FPN were excluded); (ii) being on a cortical gyrus and not on a sulcus (i.e., overlap with GM); (iii) representing the shortest perpendicular path between scalp and cortex. The entire procedure is summarized in Figure 1.

**Figure 1.**
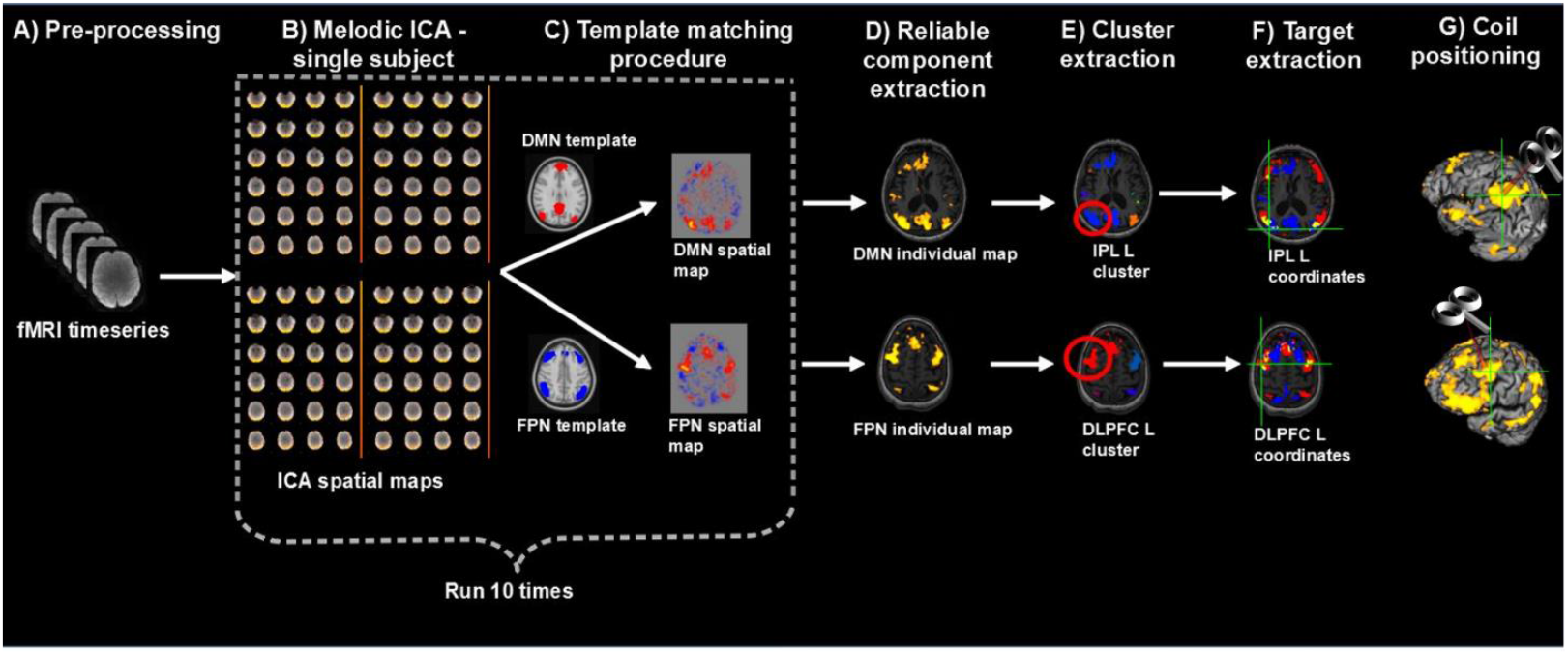
Overview of the procedure for the identification and selection of individual DMN and FPN targets based on rs-fMRI; **A)** Collected rs-fMRI data were pre-processed removing the first ten time-points, correcting motion and susceptibility-induced distortions; **B)** DMN and FPN were extracted from individual rs-fMRI scans using independent component analysis (ICA); **C)** Networks of interest (in MNI space) were identified using a template matching procedure; steps B-C were repeated 10 times; **D)** The most reliable components were identified and back-transformed to subjects’ native T1 space; **E)** Each network was decomposed into clusters and the largest cluster in the left IPL and left DLPFC was identified, for the DMN and FPN respectively; **F)** the peaks (local maxima) within these clusters were extracted and the final individual TMS targets were selected according to the following criteria: (i) location specific to the network of interest, i.e. coordinates falling within the spatial maps of both DMN and FPN (yellow areas) were excluded (blue=DMN, red=FPN); (ii) being on a cortical gyrus and not on a sulcus (i.e., overlap with GM); (iii) representing the shortest perpendicular path between scalp and cortex; **G)** TMS coil was positioned through a neuronavigation system to target the selected DMN and FPN coordinates.

The anatomical atlas label (AAL; Tzourio-Mazoyer et al., 2002) was used to label individual IPL coordinates with the corresponding anatomical region and check for anatomical-functional correspondence.

The distance between individual rs-fMRI derived and traditional anatomical coordinates was computed as follows. Individual coordinates in native space were transformed to MNI space using the affine and non-linear warping estimated by Melodic. The Euclidean distance was used to compute the distance from group-level left IPL and DLPFC coordinates reported in previous TMS studies (Cotelli et al., 2012, 2010; Fox et al., 2013; Herwig et al., 2003). Coordinates in Talairach space were transformed to MNI space using a non-linear transformation (Lacadie et al., 2008). For studies using the Brett or Lancaster transformation to derive Talairach coordinates, we used the inverse Brett/Lancaster transformation to obtain the original MNI coordinates. One-sample Wilcoxon test was used to assess whether the distance between individual and traditional coordinates exceeded two threshold’s levels, assuming a spatial extent of rTMS-induced activation of 12mm (conservative threshold; Fox et al., 2013) and 20mm (lenient threshold).

Finally, we compared the precision of our approach with traditional approaches testing i) the sensitivity of group-level IPL and DLPFC coordinates to DMN and FPN spatial maps, respectively (i.e., how frequently group-level coordinates fell into the expected network), and ii) the selectivity of this relationship (i.e., how frequently a coordinate falling into one network also fell into the other). Group-level coordinates were overlaid onto the individual spatial maps of the DMN and FPN before computing the above frequencies.

## 3. Results

Individual targets are shown relative to their network in Figure 2A, and their position relative to the coordinates reported in the literature is depicted in Figure 2B (all coordinates are reported in MNI space). The median distance between individual IPL coordinates was 18.87 mm (interquartile range: 15.41-25.57 mm). The median distance between individual DLPFC coordinates was 20.74 mm (interquartile range: 13.87-26.42 mm). When using the anatomical atlas label (AAL; Tzourio-Mazoyer et al., 2002) to localize our IPL coordinates, 4 out of 8 cases corresponded to or were close to the angular gyrus, 2 to the middle occipital gyrus, one to the inferior parietal gyrus, and one was borderline between the latter two regions (Table 2). The median distance between individual fMRI-derived and group-level IPL coordinates was >15mm for both the studies considered (Cotelli et al., 2012; Herwig et al., 2003). This distance exceeded rTMS focality when considering the 12 mm threshold (both p’s<0.05) but not the 20 mm threshold (both p’s>0.08; Table 2). The sensitivity of group-level IPL coordinates to individual DMN spatial maps was 62.5% in the best case (Herwig et al., 2003) while selectivity was generally low (>60% of the coordinates falling into the DMN also fell within the FPN) (Table 3).

**Table 2.** Individual coordinates (reported in standard MNI space) of the two targets (the left IPL node of the DMN and the left DLPFC node of the FPN) obtained with the individual rs-fMRI guided approach. The corresponding anatomical region is provided based on the Anatomical atlas label (AAL), and the average distance between individual coordinates and group-level coordinates is provided. Results of one-sample Wilcoxon tests (p-values) assessing the null hypothesis that the distance between individual and group-level coordinates is below 12mm and 20mm are reported.

| DMN – left IPL |  |  |  |  | FPN – left DLPFC |  |  |  |  |  |  |
| --- | --- | --- | --- | --- | --- | --- | --- | --- | --- | --- | --- |
|  | Individual<br>rs-fMRI<br>coordinates | AAL | Distance<br>(mm) from<br>P3 | Distance<br>(mm) from<br>IPL | Individual<br>rs-fMRI<br>coordinates | AAL | Distance<br>(mm) from<br>BA9 | Distance<br>(mm) from<br>BA46 | Distance<br>(mm) from<br>5 cm rule | Distance<br>(mm) from<br>F3 | Distance<br>(mm) from<br>BA8/9 |
| Pt 1 | -34 -80 44 | IPL | 16.16 | 27.93 | -52 24 40 | MFG | 21.26 | 24.04 | 18.60 | 16.16 | 20.10 |
| Pt 2 | -38 -66 42 | AG | 9.00 | 14.00 | -48 4 56 | PCG | 39.45 | 48.44 | 13.64 | 24.60 | 22.00 |
| Pt 3 | -44 -68 24 | MOG | 27.73 | 26.08 | -46 38 22 | MFG | 20.59 | 4.69 | 38.13 | 30.15 | 35.44 |
| Pt 4 | -52 -72 26 | AG | 29.27 | 28.07 | -48 32 32 | MFG | 15.62 | 12.08 | 27.17 | 20.12 | 25.38 |
| Pt 5 | -36 -82 42 | MOG | 18.47 | 29.39 | -58 16 40 | MFG/PCG | 31.11 | 32.34 | 20.83 | 23.35 | 25.77 |
| Pt 6 | -34 -82 44 | IPL/MOG | 17.92 | 29.80 | -50 32 38 | MFG | 15.36 | 16.91 | 23.79 | 17.12 | 22.18 |
| Pt 7 | -40 -66 36 | AG | 15.13 | 16.12 | -50 16 46 | MFG | 26.76 | 33.20 | 10.86 | 15.13 | 16.37 |
| Pt 8 | -56 -54 28 | AG | 31.58 | 21.63 | -40 28 56 | MFG | 19.29 | 33.85 | 13.64 | 8.77 | 8.25 |
| Median<br>(IQR) | **18.87**<br>**(15.41-**<br>**25.57)** |  | **18.20**<br>**(15.64-**<br>**28.50)** | **27.01**<br>**(18.88-**<br>**28.73)** | **20.74**<br>**(13.87-**<br>**26.42)** |  | **20.93**<br>**(17.46-**<br>**28.94)** | **28.19**<br>**(14.49-**<br>**33.53)** | **19.72**<br>**(13.64-**<br>**25.48)** | **18.62**<br>**(15.64-**<br>**23.98)** | **22.09**<br>**(18.24-**<br>**25.58)** |
| *p* (12 mm<br>threshold) | <0.001* |  | 0.016* | 0.008* | <0.001* |  | 0.008* | 0.039* | 0.021* | 0.023* | 0.016* |
| *p* (20 mm<br>threshold) | 0.945 |  | 0.945 | 0.078 | 0.531 |  | 0.383 | 0.383 | 1.000 | 0.844 | 0.383 |

**Table 3.** Correspondence between group-level IPL and DLPFC coordinates and individual DMN and FPN maps. As a reference, individual fMRI-derived coordinates have a sensitivity and selectivity of 100%.

| Group-level coordinates | Sensitivity | Selectivity |
| --- | --- | --- |
| **DMN** |  |  |
| P3 (Herwig et al., 2003) | 62.5% | 40% |
| IPL (Cotelli et al., 2012) | 25% | 0% |
| **FPN** |  |  |
| F3 (Herwig et al., 2003) | 75% | 50% |
| DLPFC (Cotelli et al., 2010) | 62.5% | 20% |
| 5-cm rule (Fox et al., 2013) | 62.5% | 80% |
| BA9 (Fox et al., 2013) | 25% | 0% |
| BA46 (Fox et al., 2013) | 12.5% | 100% |

**Figure 2.**
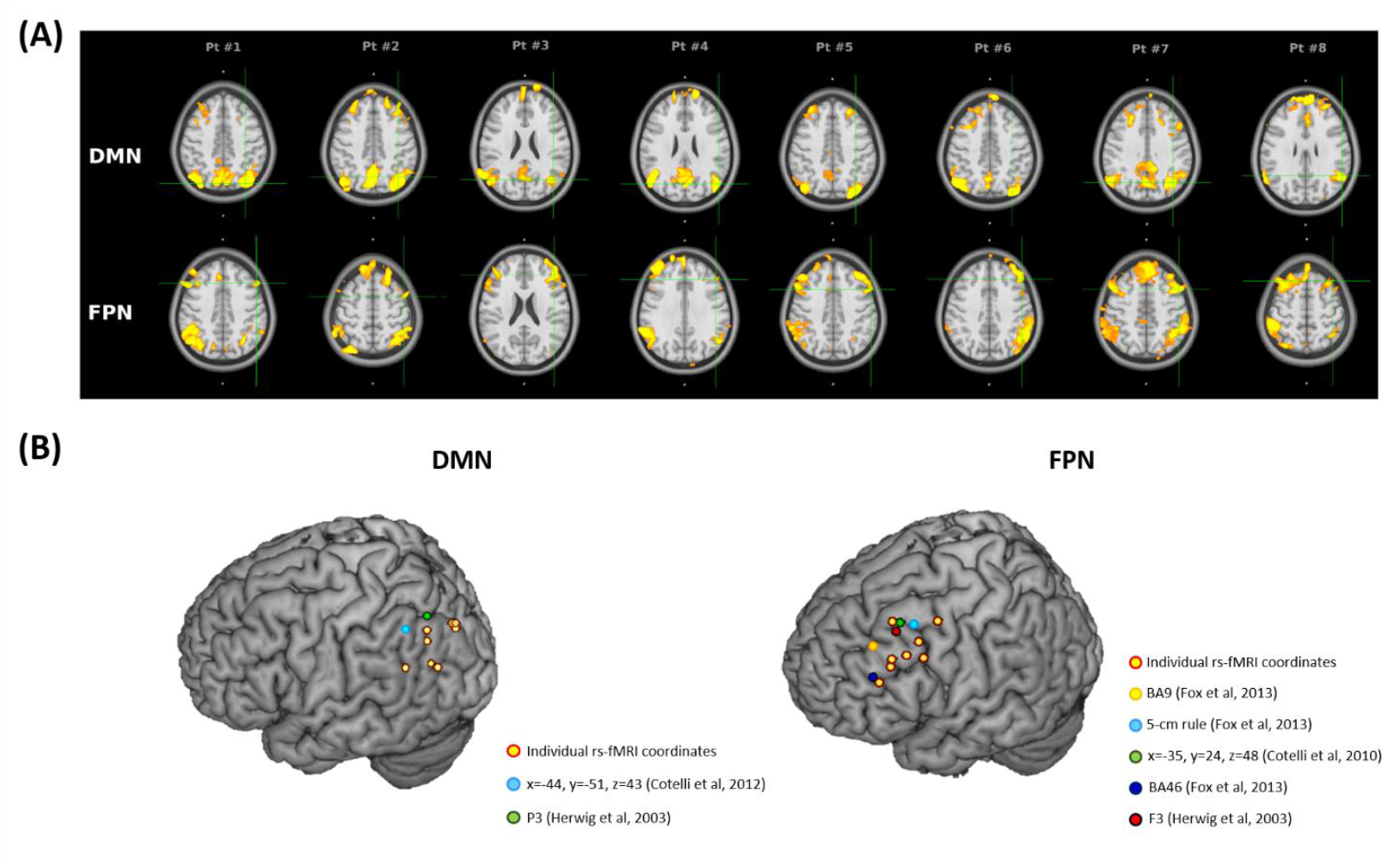
**A)** Location of the targets (reported in standard MNI space) for default mode network (DMN) stimulation (top panel) and frontoparietal network (FPN) stimulation (bottom panel) in eight AD patients. Targets (green cross) were extracted from each subject’s 3T rs-fMRI data using ICA. The DMN targets correspond to the left IPL cluster, the FPN targets to the left DLPFC cluster. The individual DMN and FPN maps are shown in orange-yellow. The targets were defined in subjects’ native T1 space and back-transformed to the standard MNI space for computation and visualization purposes; **B)** 3D render showing the individual targets (red-yellow) overlaid onto the standard MNI template. For the DMN, green target corresponds to P3 (Herwig et al., 2003), and light-blue to IPL (Cotelli et al., 2012). For the FPN, yellow target corresponds to DLPFC BA9 (Fox et al., 2013), light-blue to DLPFC-5cm rule (Fox et al., 2013), blue to DLPFC BA46 (Fox et al., 2013), red to F3 (Herwig et al., 2003), green to DLPFC BA8/9 (Cotelli et al., 2010). Key: IPL, inferior parietal lobule; AG, angular gyrus; SPG, superior parietal gyrus; MOG, middle occipital gyrus; MTG, middle temporal gyrus; PCG, precentral gyrus; MFG, middle frontal gyrus.

DLPFC coordinates were localized in the middle frontal gyrus in 6 out of 8 cases (75% of cases), in the precentral gyrus in one case, and borderline in one case. The distance between individual fMRI-derived and group-level DLPFC coordinates (Cotelli et al., 2010; Fox et al., 2013; Herwig et al., 2003) was significant for all coordinates at the 12 mm threshold (all p’s<0.05), but not at the 20mm threshold (all p’s>0.38; Table 2). Group-level DLPFC coordinates most sensitive to FPN spatial maps were F3 coordinates (75% of coordinates falling into the FPN), followed by DLPFC BA8/9 (62.5%), and 5 cm-rule (62.5%) coordinates. The selectivity of these coordinates however was high only for the 5cm rule (80% of the coordinates being specific for the FPN) and low for F3 and DLPFC BA8/9 coordinates (50% and 80% of cases also falling into the DMN, respectively) (Table 3).

## 4. Discussion

The combination of neuroimaging and neurostimulation techniques to design personalized protocols is an emerging research field, which may enhance the precision of rTMS (Cocchi and Zalesky, 2018). Here, we showed the feasibility of a tailored rTMS protocol that allows to target AD relevant networks by extracting their hub coordinates from individual rs-fMRI.

The advantages of the proposed method over previous approaches become clear when the spatial extent of TMS-induced activation is considered. Although TMS focality is difficult to estimate because of technical and anatomical factors (Thielscher and Kammer, 2004), computational models (Fox et al., 2013) indicate a physiological response to TMS within a spatial extent of 12mm when considering the mostly used standard figure-of-eight coil. Our comparisons revealed a significant distance between functionally-defined individual targets and anatomical group-level coordinates when assuming a stimulation field size <12 mm, thus favoring the spatial selectivity of our approach.

This advantage is even more striking in the hypothesis that rTMS focality is <2mm, as suggested by a recent study recording single-unit activity in the parietal cortex of rhesus monkeys (Romero et al., 2019). Even assuming a larger (e.g.., 20 mm) focality for TMS, the proposed approach has important advantages. While at a 20 mm threshold the distance between individual and traditional coordinates might not exceed TMS focality, we observed a loss of precision in targeting. Indeed, the sensitivity of group-level coordinates was 63-75% at most, indicating that in 25-33% of cases other networks will be stimulated. Moreover, the selectivity of group-level coordinates was generally low, indicating that group-level coordinates would result in stimulation of both networks rather than in the selective targeting of the intended network. The best trade-off between sensitivity/selectivity was provided by the 5 cm rule for the DLPFC node (63-80%), however these values are still less precise than our approach, which was designed to provide a sensitivity/selectivity of 100%.

The large variability observed between subjects’ spatial maps (see in particular variability between FPN maps in Figure 2A) and across individual targets (around 19-21 mm; see Figure 2B) is consistent with the knowledge that the brain’s structure and function undergo substantial changes both in physiological ageing and in AD, with a massive networks’ reorganization (Dubovik et al., 2013; Edde et al., 2020; He et al., 2020; Pläschke et al., 2020). Bearing this in mind, going beyond an anatomical approach appears crucial to increase rTMS clinical efficacy. In our sample, the functional targets did not correspond to the expected anatomical region in 25-33% of cases, confirming a lack of function-anatomical correspondence. Consistently with this view, a recent study in depression showed that the efficacy of rTMS was higher when the target was selected on the basis of functional connectivity (Weigand et al., 2018).

Notably, the proposed approach is not specific for a given TMS technique or protocol. Specifically, our strategy can be applied to both rTMS and theta burst stimulation techniques, and is not dependent on the type of stimulation protocol (i.e., inhibitory vs. excitatory). The choice of the type of stimulation to be delivered, while representing a key step in the design of TMS interventions, is outside the scope of this report. Here, we point out that TMS protocols for AD should take into account not only the localization of the target, but also the connectivity pattern (i.e., reduced vs. increased connectivity), the degree of pathology (i.e., affected vs. spared regions), and their interaction.

Furthermore, this approach can be easily translated to other dementias and diseases affected by network dysfunction in order to design TMS disorder-specific protocols. Neurodegenerative and psychiatric diseases characterized by emotional and behavioral deficits such as the behavioral variant of frontotemporal dementia (Zhou et al., 2010) and borderline personality disorder (Quattrini et al., 2019) might benefit from stimulation of the DMN and salience network, while conditions characterized by language disturbances such as primary progressive aphasia may be suited for stimulation of the language network (Ficek et al., 2019), whereas motor disorders such as Parkinson’s disease may benefit from stimulation of the sensorimotor network (Göttlich et al., 2013).

Some possible limitations of the proposed approach should be mentioned. To be clinically usable, individualized coordinate extraction from rs-fMRI needs to be reliable. This requires i) the definition of standard pre-processing procedure and ii) that networks are reliable. For the first issue, while our procedure is relatively straightforward, it requires independent validation. Moreover, while we used ICA, seed-correlation analysis is a valid alternative that has already been applied in other studies (Nilakantan et al., 2019). Seed-based approaches typically use the hippocampus as seed region to derive the DMN parietal node, defined as the most functionally correlated region. While we used a different strategy (based on the local cluster maxima) that does not provide information on the strength of the correlation with the hippocampus or other DMN regions, our approach extracted the region most involved and active within the DMN component. Moreover, one advantage of ICA-based compared to seed-based approaches is that they enable to extract statistically independent sources, while the latter cannot distinguish whether a brain region is shared by multiple networks. Furthermore, in our study we used advanced fMRI sequences (multiband, 600 volumes), which may not be available at all clinical centers. For the second aspect, in our study we counterbalanced this issue by extracting the network 10-fold and ensuring that the same component was extracted reliably. Several automated tools are available to assess networks reliability (e.g., ICASSO; Himberg et al., 2004) and the use of these tools is recommended to ensure that the extracted networks are stable enough for rTMS targeting. Finally, while we expect that our approach would increase rTMS efficacy by increasing the precision of target localization, this was not formally tested. Forthcoming studies testing the differential impact of network-based versus traditional approaches on clinical outcomes are needed to directly test this assumption.

In conclusion, based on a functional network perspective, we proposed a procedure for individual identification of TMS targets, paving the way for unprecedented personalized connectivity-based rTMS treatments for AD.

## Data Availability

Chiara Bagattini, Debora Brignani, Sonia Bonni', Roberto Gasparotti, Michela Pievani (2020): rsfMRI network-based TMS targeting, Mendeley Data, V1

http://dx.doi.org/10.17632/5zxyrvc5nz.1

## Acknowledgements

This work was supported by a grant from the Italian Ministry of Health awarded to DB, SB and MP (Bando Ricerca Finalizzata 2016 – grant number: GR-2016-02364718). The funder had no role in study design, data collection and analysis, decision to publish, or preparation of the manuscript.

## Data Availability

Bagattini, Chiara; Brignani, Debora; Bonnì, Sonia; Gasparotti, Roberto; Pievani, Michela (2020), “rs-fMRI network-based TMS targeting”, Mendeley Data, V1, doi: 10.17632/5zxyrvc5nz.1

